# Do Pandemics Obey the Elliott Wave Principle of Financial Markets?

**DOI:** 10.1101/2021.01.21.21250273

**Authors:** Prashant Dogra, Eugene J. Koay, Zhihui Wang, Farhaan S. Vahidy, Mauro Ferrari, Renata Pasqualini, Wadih Arap, Marc L. Boom, H. Dirk Sostman, Vittorio Cristini

**Affiliations:** Mathematics in Medicine Program, Houston Methodist Research Institute, Houston, Texas, USA; Department of Gastrointestinal Radiation Oncology, The University of Texas MD Anderson Cancer Center, Houston, TX, USA; Center for Outcomes Research, Houston Methodist Research Institute, Houston, Texas; Houston Methodist Neurological Institute, Houston Methodist, Houston, Texas; Department of Pharmaceutics, University of Washington, Seattle, Washington, USA; Dompé X-Therapeutics, San Mateo, California, USA; Rutgers Cancer Institute of New Jersey, Newark, NJ, USA; Department of Radiation Oncology, Division of Cancer Biology, Rutgers New Jersey Medical School, Newark, NJ, USA; Department of Medicine, Division of Hematology/Oncology, Rutgers New Jersey Medical School, Newark, NJ, USA; Department of Medicine, Houston Methodist, Houston, Texas; Weill Cornell Medicine, New York, NY, USA; Houston Methodist Research Institute, Houston, TX, USA; Houston Methodist Academic Institute, Houston, TX, USA

## Abstract

The Elliott Wave principle is a time-honored, oft-used method for predicting variations in the financial markets. It is based on the notion that human emotions drive financial decisions. In the fight against COVID-19, human emotions are similarly decisive, for instance in that they determine one’s willingness to be vaccinated, and/or to follow preventive measures including the wearing of masks, the application of social distancing protocols, and frequent handwashing. On this basis, we postulated that the Elliott Wave Principle may similarly be used to predict the future evolution of the COVID-19 pandemic. We demonstrated that this method reproduces the data pattern especially well for USA (daily new cases). Potential scenarios were then extrapolated, from the best-case corresponding to a rapid, full vaccination of the population, to the utterly disastrous case of slow vaccination, and poor adherence to preventive protocols.

---

In 1938, Ralph Nelson Elliott, an accountant by profession, argued that the stock market is a phenomenon governed by crowd psychology^1^. In his classic books *The Wave Principle*^1^ and *Nature’s Law: The Secret of the Universe*^2^, he described that human emotions, which tend to be rhythmical and follow patterns of optimism and pessimism, are instrumental in shaping market dynamics. The sentiments of the crowd can lead to quantifiable patterns in human activities, and when applied to financial markets, such patterns are observed in the movement of stock and commodity prices across time scales ranging from hours to years. These observations made by Elliott lead to his description of the Wave Principle, which is a phenomenological principle used by traders to understand market behavior and predict future trends. Given the ability of Elliott Wave Principle to describe human sentiments, we hereby hypothesized that the Wave Principle could also describe the ongoing COVID-19 pandemic.

## Elliott Wave Principle basics

In brief, the Wave Principle states that movement of stock and commodity prices occurs on a wave, where a completed movement is made up of five sub-waves. As shown in **Figure 1**, sub-waves i, iii, and v (black lines) are in the direction of movement (known as *impulses*), while sub-waves ii and iv (black lines) are contrary to the direction of movement (known as *correctives*). Elliott identified a fractal character in the waves of stock and commodity prices, such that the five sub-waves of one dimension or degree (i.e., sub-waves i, ii, iii, iv, v) become the first wave of the next higher dimension (i.e., wave 1), and so on. Notably, impulses and correctives differ from each other such that the former are divisible into five sub-waves of lesser dimension (i to v), while the latter are divisible into three sub-waves of lesser dimension (a, b, c). The impulsive and corrective moves of an Elliott wave follow *Fibonacci ratios*^3^, which are mathematical relationships between the numbers of the Fibonacci sequence^1^. The various waves of an Elliot wave relate to each other through Fibonacci ratios and thus the ratios are used to predict the extensions and retracements in stock and commodity prices, a technique known as the *Fibonacci Pinball*^4^. Since the wave principle is based on sentiments of the crowd, we reasoned that other fields of human activity involving masses might perhaps behave in a fashion predictable by the Wave Principle.

**Figure 1.**
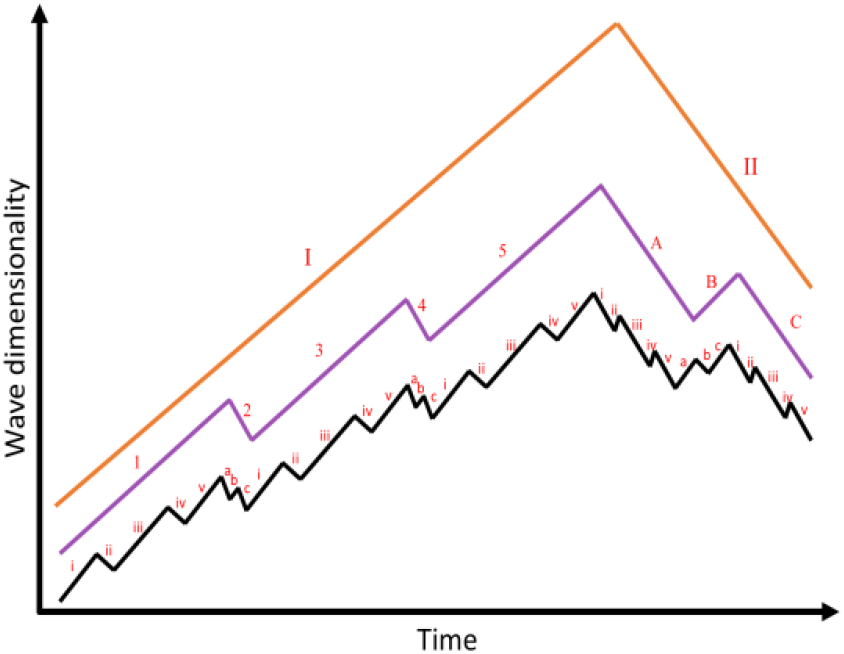
Elliott waves of smallest dimension (sub-waves; black) are shown along with the waves of higher dimensions (purple and orange). i, iii, v are impulses of the smallest dimension; 1, 3, 5 are impulses of next higher dimension; I is the impulse of next higher dimension. Similarly, ii and iv are correctives of the smallest dimension, 2 and 4 are correctives of the next higher dimension, and II is the corrective of next higher dimension. Impulses (e.g. I) are made up of 5 waves of lesser dimension (i.e. 1, 2, 3, 4, 5), while correctives (e.g. II) are made up of 3 waves of lesser dimension (i.e. A, B, C).

## Applying the Wave Principle to the COVID-19 pandemic

Given that more than 86 million people have been infected and more than 1.87 million patients have died worldwide due to COVID-19 as of January 2021^5^, human emotions have thus been running particularly high during the pandemic. Furthermore, especially with the shutdown of economies and social isolation due to travel restrictions and lockdowns, human emotions have been going back and forth while adjusting to “the new normal.” Against this backdrop, we have looked at the seven-day moving average of daily new cases of COVID-19 disease, in hard-hit countries/regions, since the beginning of the pandemic^6^.

Indeed, as shown in **Figure 2a**, the trend for USA seems to be obeying the Wave Principle on a timescale of weeks, i.e. it exhibits an initial impulse (wave 1), followed by a corrective (wave 2), and then an extended impulsive wave 3, which is made up of 5 sub-waves of smaller dimension (i.e., sub-waves i, ii, iii, iv, and v). The current trend seems to be heading towards a corrective wave 4, which is comprised of sub-waves a and b (c is yet to show). This pattern can be attributed to crowd psychology that manifested itself in the form of a rhythmical pattern of adherence to public health policies imposed by the government, due to fear of infection, followed by either relaxation of policies by the government, or disobedience to policies by the masses, due to economic turmoil and growing resentment. As currently evident, the imposition of international travel restrictions, stay-at-home orders, and closure of schools and businesses due to growing number of cases around mid-March 2020, followed by mandatory use of masks in public places starting mid-July 2020, largely promoted the corrective waves (during April and May 2020 (wave 2) and August 2020 (wave ii), respectively). However, these correctives may have given the masses a false sense of the infection “being over with” and leading into them becoming less vigilant. Also, the growing unrest in public due to closure of businesses and plummeting GDP in the second quarter of 2020 has led to relaxation of some policies by the government, particularly opening of businesses like restaurants, bars, and gyms in early June 2020, and then schools in early September 2020, thereby instigating the impulses thereafter (wave i in late June and July 2020 and wave iii in October and November 2020, respectively). Of note, the third wave of an Elliott wave is typically the largest and the most powerful, which is also evident above in the observation that wave 3 has brought about an increase in daily cases by one order of magnitude compared to the value at the base of wave 2, thereby adhering to the wave principle. Similarly, the smaller dimensional sub-wave iii is also the strongest impulse of wave 3.

**Figure 2.**
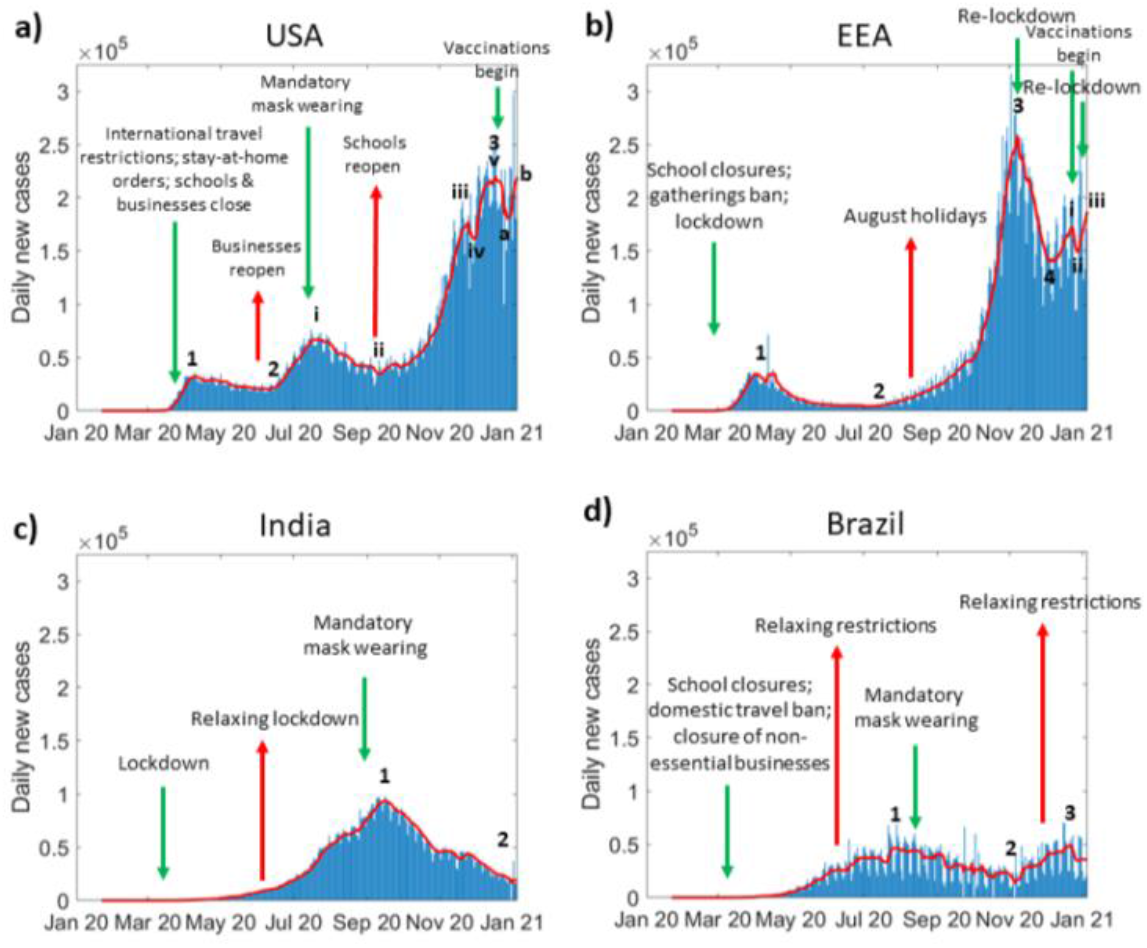
Daily new case timeline of COVID-19 in **a**) USA, **b**) European Economic Area (EEA), **c**) India, and **d**) Brazil, as of 1/5/2021. Red line represents seven-day moving average of the cases. Impulses 1, 3, 5, (or, i, iii, v) and correctives 2 and 4 (or, ii and iv) are shown wherever they are visible. Green and red arrows indicate the major restrictive and permissive events, respectively. Data source: *https://github.com/owid/covid-19-data/blob/master/public/data/jhu/new_cases.csv*

Many other countries seem to be obeying the principle, although none seem to have completed super-wave I (higher dimension) yet. For instance, after an initial impulsive wave 1 (**Figure 2b**), the European Economic Area (EEA, comprising of 32 European countries) had an extended corrective wave 2 following stringent restrictions starting March 2020 that lasted through summer 2020. The extended corrective wave must have made the crowds less watchful during the European holiday season in August 2020, following which a strong impulsive wave 3 was seen, which was partially corrected through wave 4 by renewed restrictions starting mid-October in most European nations. An uptick around mid-November 2020 is indicative of the beginning of impulsive wave 5, comprised of sub-waves i, ii, iii (iv and v are yet to show). Further, India (**Figure 2c**) and Brazil (**Figure 2d**), the other two most hard-hit countries, imposed restrictions early-on in the pandemic during March 2020 that lasted around eight weeks, which helped delay impulsive wave 1 in these countries. But, after the easing of restrictions in June 2020 due to declining economy and growing unrest in the masses, impulsive wave 1 took off. While no new nation-wide restrictions were then imposed, but due to other measures like mandatory use of masks in public places and suggestive social distancing, a corrective wave 2 followed. While India continues to be on the corrective wave 2, Brazil is exhibiting an impulsive wave 3 that can be attributed to the revival of tourist attractions, beaches, shopping malls, and restaurants since October 2020^7^. It is important to note that in case of India, there is growing evidence that the Indian population possesses better innate immunity to SARS-CoV-2^8-10^, which indicates that any possible divergence from the sentiment-mediated wave principle may be a result of the additional biological variable at play in this population.

## The “real” crisis may happen in 2021

Since mid-December 2020, vaccines are starting to become available hierarchically, based on the susceptibility of populations, in high-income countries. However, due to logistical limitations, vaccines may not be available to the larger population anytime soon^11, 12^, but irrespective of that, based on a survey published by Pew Research Center, *39% of the US adults have expressed lack of intent to get a vaccine, and 62% express being uncomfortable to be the first ones to get the vaccine*^13^. On the other hand, the majority of the population (∼80%) in upper-(e.g. Brazil) and lower-middle (e.g. India) income countries may not receive the vaccine until the end of 2021^14^. Moreover, the recent emergence of variants of SARS-CoV-2 in the UK and South Africa, capable of increasing the reproduction number (R0) of COVID-19, are concerning for their increased infectivity and transmissibility and the potential to cause vaccine failure^15-17^. While SARS-CoV-2 mutates less frequently than influenza viruses, and breakthrough infections due to vaccine failure have not yet been registered, but mutations over time may require alterations in vaccines, thereby slowing down the attainment of herd immunity^18^. Thus, for the above reasons, one may conjecture that if effective vaccines are not administered globally to the masses anytime soon, the long rally of new infections will complete impulse I of the higher dimensional wave by mid-2021, which will inevitably exhibit waves II, III, IV, and V over the coming year or two. This will potentially lead to a massive rise in new infections and deaths, unless the socio-medical parameters are changed by permanent adherence to stringent policies *and* introduction of vaccines readily available to the larger population.

By using the Fibonacci Pinball method of Elliott Wave analysis^3, 19^, we can make projections for how the daily cases will evolve over time in the coming year. As shown in **Figure 3**, with USA as the reference example, the retracements (R) and extensions (E) assumed by the waves thus far approximately adhere to Fibonacci ratios, thereby suggesting that the pandemic has adhered to the Wave Principle and it is reasonable to use Fibonacci Pinball to make future projections. Since the current wave of infections (sub-wave b of wave 4) seems to have peaked, it will be followed by a corrective sub-wave c of wave 4, and then an impulsive wave 5 to complete the higher dimensional super-wave I. We show the worst-, one of the intermediates-, and best-case scenarios, using 23.6% E and 261.8% E (dashed red line), 100% E and 23.6% E (dashed-dotted red line), and 500% E and 0.69% E (dotted red line), respectively for the evolution of the pandemic over the next several months. While the (highly improbable) best case scenario predicts a strong corrective sub-wave c of wave 4, followed by a negligibly impulsive wave 5 (primarily due to availability of vaccines to the masses and stringent nation-wide public health policies in place), leading to ∼90% reduction in new daily infections by early-fall 2021. Contrarily, the worst-case scenario (more probable due to unavailability of vaccines to the masses, reopening of schools and businesses, and spread of highly transmissible mutant strains due to limited international travel restrictions) predicts a weak corrective sub-wave c of wave 4 and a strong impulsive wave 5, leading to a daily new infection count of ∼720,000 by early-fall 2021. Alternatively, an intermediate-scenario can be predicted with mediocre strengths of correctives and impulses, which leads to a daily case count in early-fall 2021 that is comparable to the current daily count, mainly occurring because of partial availability of vaccines and limited restrictions imposed. Either way, the corrective sub-wave c of wave 4, especially if deep, will give the public the illusion that the pandemic is fading away, thus leading to relaxation of restrictions and fueling impulsive wave 5.

**Figure 3.**
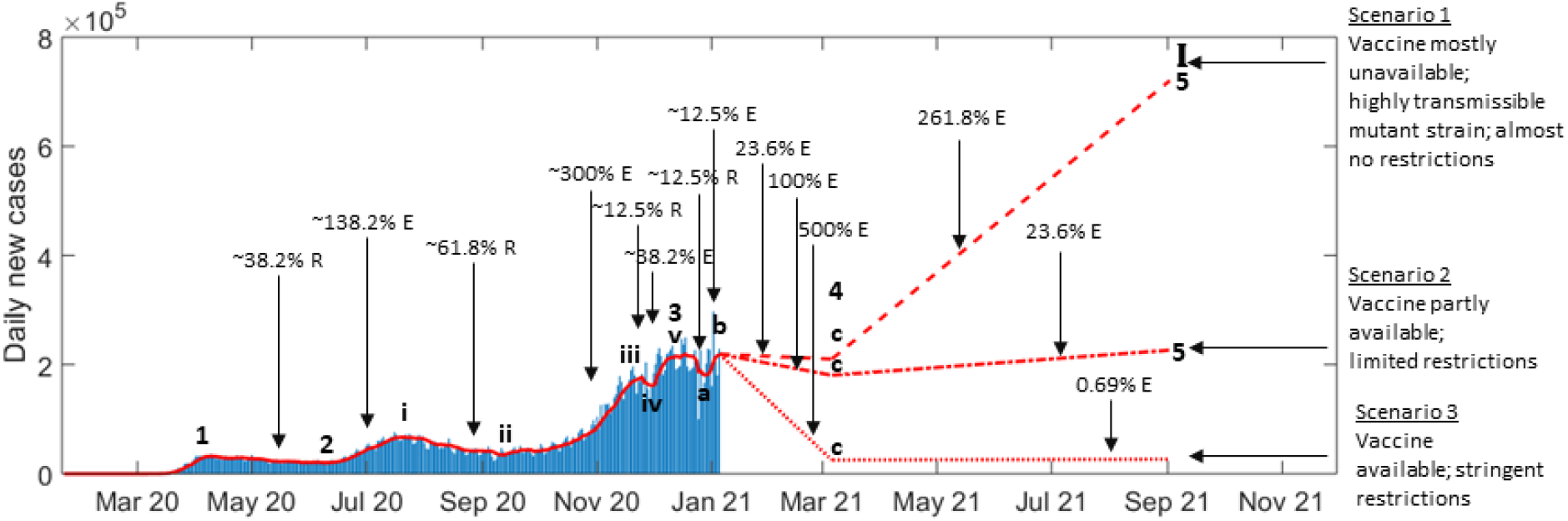
Elliott wave analysis for USA using the Fibonacci Pinball method. Dotted, dashed-dotted, and dashed red lines show predictions of best-, intermediate-, and worst-case scenarios, respectively, for daily new infections. Black arrows point out the closest Fibonacci ratio exhibited by an extension (E) or a retracement (R) of the wave.

Thus, as shown in **Figure 3**, if our estimates are correct, then the *real crisis of this pandemic is yet to arrive in countries that have lax COVID-19 policies*. As a representative example, if current policies continue in the United States as they did in 2020, wave 5 will likely last the better part of 2021 and affect the vast majority of the US population (note that this is typical for such one-in-a-century pandemic events). While the Biden administration is expected to change current federal approaches to the pandemic, adoption of these policies uniformly throughout the United States is expected to be challenging due to sociopolitical resistance. In the absence of a widely distributed and effective vaccine *as well as* stringent public health policies throughout, the worst of the pandemic for the United States is yet to come.

While China was presumably the first place to be hit by COVID-19, but regardless it was able to control the infection swiftly and efficiently due to the existence of a centralized epidemic response system, which also gained support from the masses due to the mental preparedness of adults who had witnessed the fatality of SARS-CoV epidemic in 2002-2003^20^. It is thus evident that human emotions act as the fuel for the fury of a pandemic, and thus without *both* stringent public health policies and availability of a vaccine to the larger population to achieve herd immunity, it will be virtually impossible to control the pandemic. Thus, it will not be wrong to speculate that the growing pandemic fatigue among the masses will make the public lower its guard against SARS-CoV-2, and the additional absence of a vaccine is a recipe for disaster that is waiting to unfold itself to extend the pandemic deep into 2021. Historically, the H1N1 influenza pandemic (also called Spanish flu) of 1918-1920, which is one of the most severe pandemics of history, occurred in three major cycles of illness^21^ and caused deaths of ∼50 million people globally, with 675,000 in the United States alone^22^. Since the case-fatality ratio of Spanish flu was ∼2.5% and the population of USA in 1918 was ∼103 million, therefore ∼27 million Americans must have contracted the infection^23^. Assuming that the same fraction of population will be infected by COVID-19, it can be estimated that potentially *∼85 million Americans will be infected by the pandemic, and with a case-fatality ratio of 1.8% in the USA*^*24*^, *∼1.5 million deaths can occur*. Note that with the availability of more FDA-approved therapeutics for COVID-19, the case-fatality ratio is expected to go down further, thereby decreasing fatalities. The three cycles of Spanish flu deaths are equivalent to one complete movement of the Elliott wave, i.e. the three major death events correspond to waves I, III, and V (impulses) of an Elliott wave (**Figure 4**), out of which wave III was the deadliest, thereby lending further evidence to our predictions. *Thus, to win over the COVID-19 pandemic effectively, it is critical to empower the corrective wave 4 by effective implementation of public health policies, even in the face of decreasing infections, to buy time for mass production of vaccines*.

**Figure 4.**
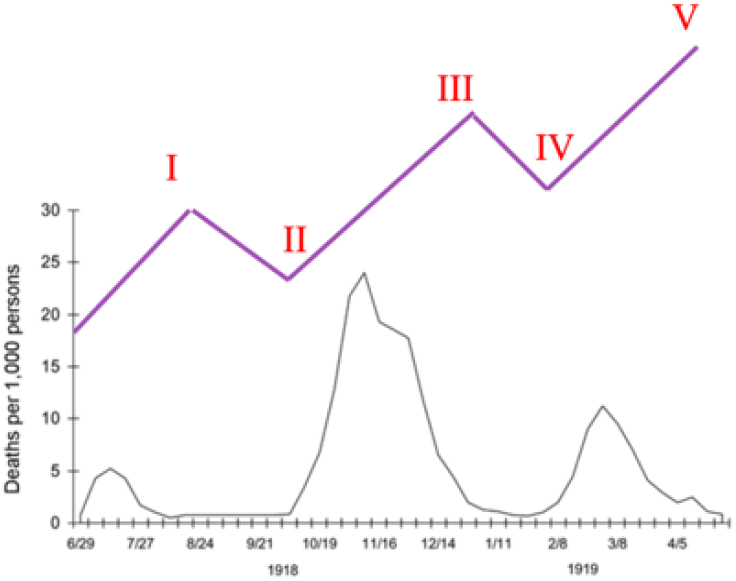
Three cycles of illness during pandemic influenza (Spanish flu) of 1918-1920 (black). Hypothetical Elliott wave (purple) is overlaid on the plot to demonstrate correspondence of deaths with different waves of the pandemic. Adapted from: *https://www.cdc.gov/flu/pandemic-resources/1918-commemoration/three-waves.htm*

## Data Availability

n/a

## Author Contributions

VC conceived the study. PD curated the data. PD and VC performed the analysis. PD, EJK, ZW, FSV, MF, RP, WA, MLB, HDS, and VC interpreted the results. PD and VC wrote the manuscript. EJK, ZW, FSV, MF, RP, WA, MLB, and HDS edited the manuscript.

## Acknowledgements

The research reported in this manuscript was supported by the National Science Foundation grant DMS-1930583 (to Z.W. and V.C.), National Institutes of Health grants 1U01CA196403 (to E.J.K., Z.W. and V.C.), 1U01CA213759 (to Z.W. and V.C.), 1R01CA226537 (to Z.W., R.P., W.A. and V.C.), 1R01CA222007 (to Z.W. and V.C.) and U54CA210181 (to E.J.K., Z.W. and V.C.). The funders had no role in study design, data collection and analysis, decision to publish or preparation of the manuscript.

## Footnotes

1 In a Fibonacci sequence, each number is the sum of the two preceding numbers. Thus, the sequence is: 0, 1, 1, 2, 3, 5, 8, 13, 21, 34, and so on.

